# Towards Predicting 30-Day Readmission among Oncology Patients: Identifying Timely and Actionable Risk Factors

**DOI:** 10.1101/2022.01.05.21268065

**Authors:** Sy Hwang, Ryan Urbanowicz, Selah Lynch, Tawnya Vernon, Kellie Bresz, Carolina Giraldo, Erin Kennedy, Max Leabhart, Troy Bleacher, Michael R. Ripchinski, Danielle L. Mowery, Randall A. Oyer

## Abstract

1

**Purpose:** Predicting 30-day readmission risk is paramount to improving the quality of patient care. Previous studies have examined clinical risk factors associated with hospital readmissions. In this study, we compare sets of patient, provider, and community-level variables that are available at two different points of a patient’s inpatient encounter (first 48 hours and the full encounter) to train readmission prediction models in order to identify and target appropriate actionable interventions that can potentially reduce avoidable readmissions.

**Methods:** Using EHR data from a retrospective cohort of 2460 oncology patients, two sets of binary classification models predicting 30-day readmission were developed; one trained on variables that are available within the first 48 hours of admission and another trained on data from the entire hospital encounter. A comprehensive machine learning analysis pipeline was leveraged including preprocessing and feature transformation, feature importance and selection, machine learning modeling, and post-analysis.

**Results:** Leveraging all features, the LGB (Light Gradient Boosting Machine) model produced higher, but comparable performance: (AUROC: 0.711 and APS: 0.225) compared to Epic (AUROC: 0.697 and APS: 0.221). Given features in the first 48-hours, the RF (Random Forest) model produces higher AUROC (0.684), but lower AUPRC (0.18) and APS (0.184) than the Epic model (AUROC: 0.676). In terms of the characteristics of patients flagged by these models, both the full (LGB) and 48-hour (RF) feature models were highly sensitive in flagging more patients than the Epic models. Both models flagged patients with a similar distribution of race and sex; however, our LGB and random forest models more inclusive flagging more patients among younger age groups. The Epic models were more sensitive to identifying patients with an average lower zip income. Our 48-hour models were powered by novel features at various levels: patient (weight change over 365 days, depression symptoms, laboratory values, cancer type), provider (winter discharge, hospital admission type), community (zip income, marital status of partner).

**Conclusion:** We demonstrated that we could develop and validate models comparable to existing Epic 30-day readmission models, but provide several actionable insights that could create service interventions deployed by the case management or discharge planning teams that may decrease readmission rates over time.

## 2 Introduction

Hospital readmissions affect patient outcomes and increase healthcare utilization and costs. Since 2012, hospitals have seen reduced Medicare payments for a subset of excess readmissions. Medicare’s value-based purchasing program encourages hospitals to improve communication and care coordination to better engage patients and care-givers in discharge plans and, in turn, reduce avoidable readmissions. In 2014, the Comprehensive Cancer Center Consortiums for Quality Improvement (C4QI) developed a 30-day readmission measure for oncology to aid hospitals in detecting and preventing potential readmissions among oncology patients.^1^ Since then, researchers and healthcare providers alike have studied risk factors to predict preventable readmissions among oncology patients.

### 2.1 Risk Factors for Predicting 30-day Readmissions

Among cancer patients, the landscape of risk factors potentially associated with 30-day readmissions is vast and diverse. Researchers have studied the relationships between demographics, clinical course, social determinants of health, and cancer types among other risk factors recorded in the electronic health record (EHR) and their role in predicting 30-day readmissions. Whitney et al. trained log-linear Poisson regression models to identify risk factors associated with higher readmission rates among advanced cancer patients. They observed significant associations with black non-Hispanic and Hispanic patients; public insurance and no insurance; lower socioeconomic status quintiles; more than one comorbidity; and pancreatic and non–small cell lung cancers.^2^ Given that age is a critical factor in readmission risk, Chiang et al. conducted a multivariate analysis of predictors for 30-day readmission among older adult cancer patients.^3^ Multivariable analyses identified dependencies in feeding and housekeeping prior to admission as associated with higher odds of readmission. Age less than 75, black race, potentially inappropriate medications, and high-risk reasons for index admission were also associated with increased odds of readmission. Furthermore, thoracic, hematologic, and gastrointestinal cancers were most common among those readmitted. Burhenn et al. built upon this work to investigate additional predictors of hospital readmission among older adults (greater than 65 years of age) with cancer.^4^ Predictors that informed their conditional logistic regression model included: patient/cancer characteristics; functional status; fall risk; chemotherapy line; comorbidities; laboratory values; discharge parameters; and miscellaneous information (do not resuscitate order, pain scores). They observed that older cancer patients with two or more abnormal laboratory results (hemoglobin, albumin, sodium, and serum glutamic oxaloacetic transaminase) at discharge were three times more likely to be readmitted within 30 days compared to those with less than two abnormal results.

### 2.2 Predicting Readmissions using Machine Learning

However, not all predictors of 30-day readmission are available at the time of discharge planning, which often begins within the first 24-to-48 hours of admission. To study the impact of temporally-available clinical features on readmission, Wong et al. trained nonparametric gradient-boosted tree models with tree-explainers based on information available within the first 24 hours versus information available close to discharge. They further enriched their feature set with clinical embeddings representing the most recent 50 International Classification of Disease (ICD) 9 diagnoses within the six months preceding admission and a time point during the hospital visit and the most recent 50 abnormal laboratory values between admission and the time point.^5^ Their most predictive model achieves an area under receiver operating characteristic curve (AUROC) of 0.78 using features available from readmission through discharge including ICD-9 billing codes and Logical Observation Identifiers Names and Codes (LOINC) embeddings, compared to an AUROC of 0.74 using features available within 24 hours after admission including baseline factors and ICD-9 clinical embeddings. Among the most informative features available within the first 24 hours include: ICD-9 embeddings; number of admissions in the previous six months; age at admission; surgery; and, hematology. Additional predictive power was obtained with features available at discharge including: LOINC embeddings; length of stay; number of LOINC codes; and, number of ICD-9 codes.

Our study builds upon these works by developing predictive models that identify actionable patient-level, provider-level, and community-level insights available to care providers within the first 48 hours of admission which could be addressed to reduce the risk of readmission among cancer patients. We applied a rigorous analysis pipeline including a diversity of supervised machine learning strategies to identify the best predictive model. Additionally, we determined the most predictive features among several machine learning algorithms to emphasize the fidelity of insights learned about readmission risk factors. Our four-fold short-term goals are to: 1) compare prediction models using several machine learning algorithms to predict 30-day readmission using risk factor information available from the full visit versus first 48 hours; 2) learn the relative importance of risk factors across classifiers for predicting 30-day readmission; 3) compare our best models to existing EHR-based classifiers; and, 4) determine timely and actionable interventions to reduce the likelihood readmissions.

## 3 Methods

This study was reviewed and approved by the University of Pennsylvania (#834695) and the Lancaster General Hospital (LGH) Institute Review Boards (#2019-51), respectively. The dataset was de-identified for data analysis. This study complied with the transparent reporting of a multivariable prediction model for individual prognosis or diagnosis.^6^

### 3.1 Cohort and Dataset

The study is a retrospective analysis of cohort data where outcomes are compared between patients that were readmitted and not readmitted within a span of 30 days after discharge. We queried all oncology patients at the Ann B. Barshinger Cancer Institute (ABBCI) in Lancaster, PA. Our cohort consists of 2,460 patients above the age of 18 who were active patients at ABBCI and had a cancer diagnosis documented within the top 3 of their diagnoses list in the EHR within a 3-year time frame between July 2016 and July 2019. Patients who were known to have moved away from the area, expired during the 30-day readmission window, or had a readmission at a different hospital were removed from the study. Based on a list of all hospital encounters within the selected cohort time frame, patients who experienced a readmission within a 30-day window after discharge were given the positive class label while the rest of the cohort were placed in the negative class. For the positive class, the encounter immediately preceding the readmission encounter is used as input for our model. For all patients that have both readmission encounters and non-readmission encounters, they are randomly selected into either class in order to avoid any bias. Lastly, the dataset was partitioned into separate development and validation datasets for model cross-validation training and testing evaluation followed by secondary evaluation with the single hold-out validation data. The development set was comprised of encounters from July 2016 and June 2018; the validation set was comprised of encounters from July 2018 through June 2019.

### 3.2 Predictive Variables

The variables included are selected based on literature review and oncology experts at the ABBCI. Broadly, they can be categorized into three levels of insights: **patient, provider**, and **community** in **Table 1**. Patient-level types include demographics, social history, laboratory results, severity indicators, and cancer specific information. Provider-level feature types include clinical course, therapeutics administered, administrative events, and the LACE risk score index (a composite score that is the gold standard for predicting 30-day readmissions at the point of care). Community-level feature types include social determinants of health, living situation, and other socio-economic indicators.

**Table 1:**
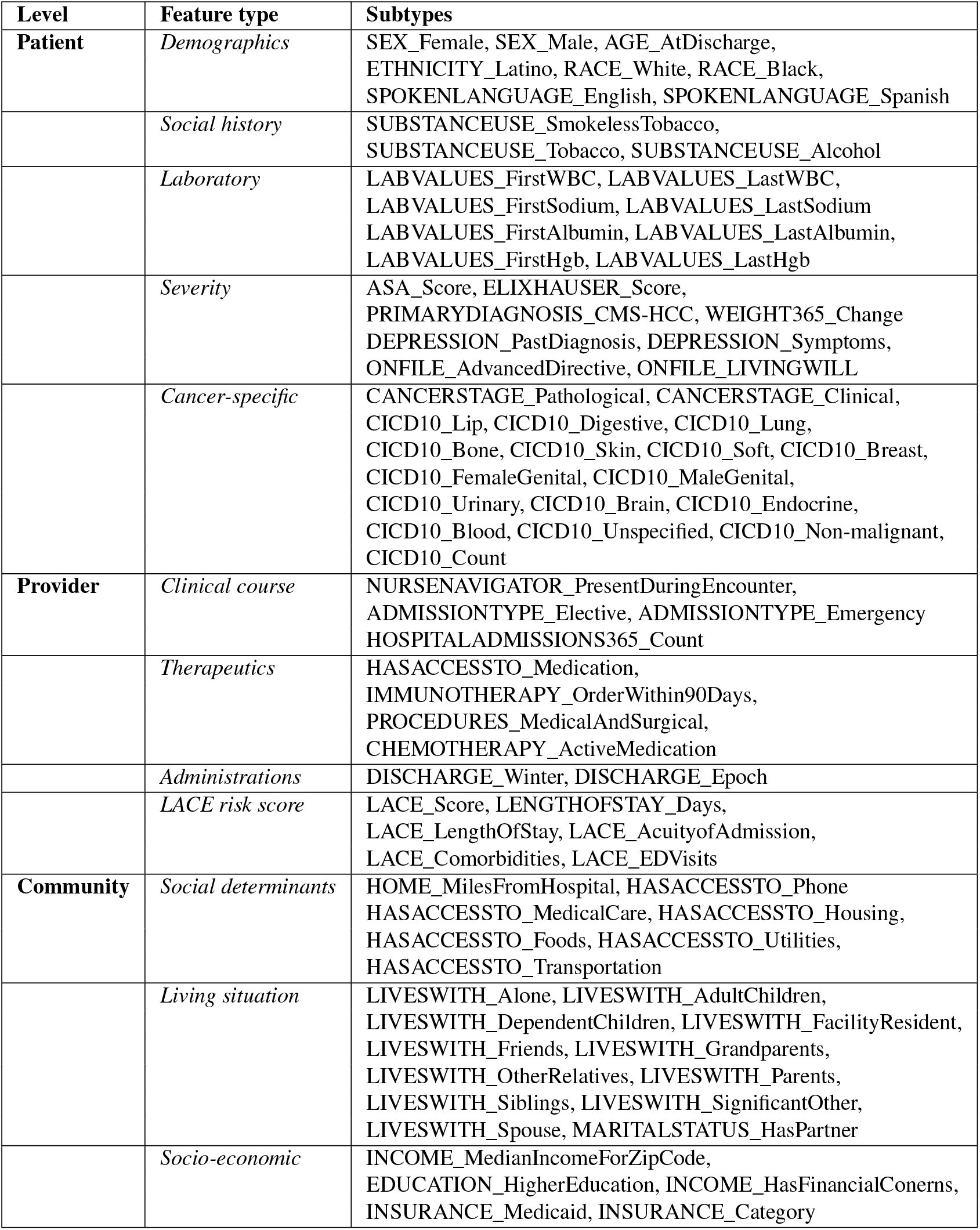
CLINICAL FEATURES and their Values. ASA = American Society of Anesthesiologists; WBC = white blood cell; Hgb = hemoglobin; CMSHCC_DX = Center for Medicare Medicaid Services Hierarchical Condition Category

### 3.3 Machine Learning Analysis Pipeline

Machine learning (ML) analyses were conducted using a recently developed rigorous ML analysis pipeline for binary classification. This pipeline was designed to compare predictive ML modeling performance and identify feature importance consensus over a variety of established ML modeling approaches implemented in or compatible with scikit-learn.^7^ The pipeline was designed to avoid common modeling biases (e.g., sample bias, and data leakage), and ensure sensitivity to complex associations with outcome including epistatic feature interactions. An early version of this pipeline was previously applied to epidemiological investigation of pancreatic cancer.^8^ For this paper, we expanded this pipeline in three ways for readmission prediction: 1) added k-nearest neighbors classification and gradient boosting for a total of 11 unique established ML modeling algorithms, 2) uniformly applied permutation-based feature importance estimation for all ML models, and 3) added the ability to automatically evaluate all trained models on a held-out validation dataset. **Figure 1** illustrates the components of this ML pipeline and we outline specific elements below. The entirety of this pipeline has been made available here: https://github.com/UrbsLab/AutoMLPipe-BC.

**Figure 1:**
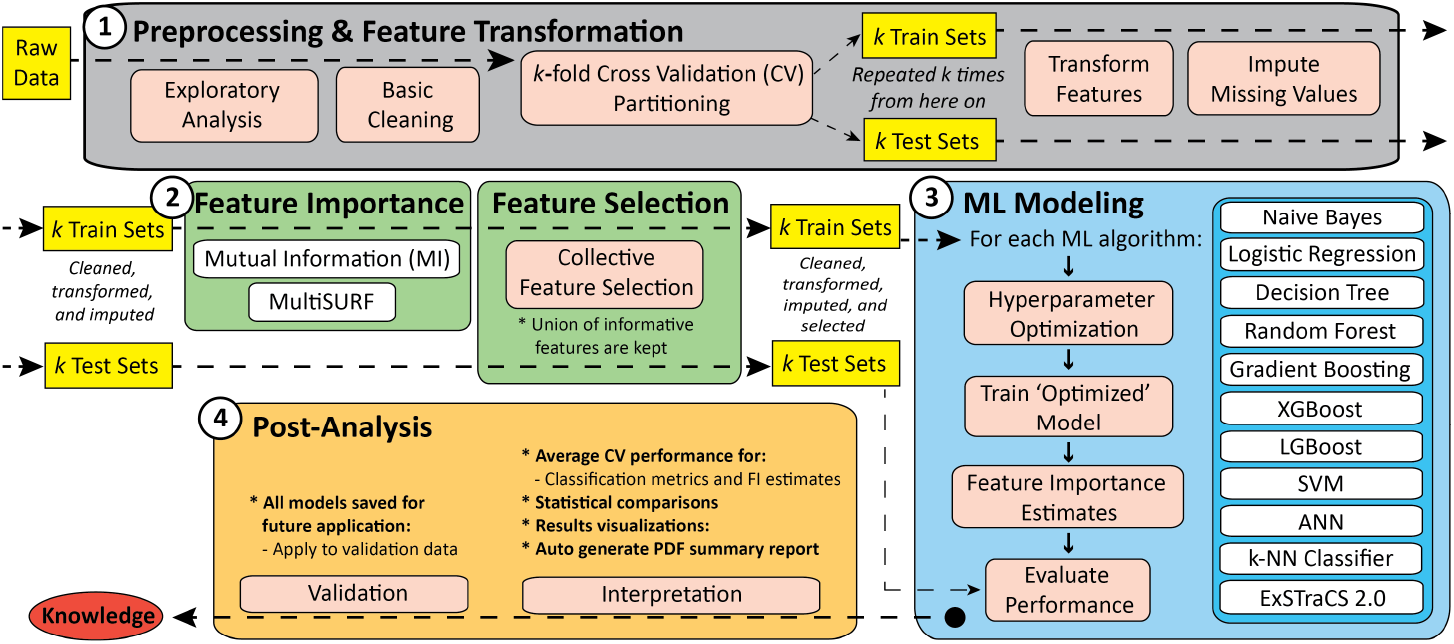
Schematic of the rigorous ML analysis pipeline applied in this study separated into four phases

#### 3.3.1 Phase 1: Preprocessing and Feature Transformation

Phase 1 of the ML pipeline first automates exploratory analysis and basic data cleaning (i.e. removing any instances that may be missing a class label, e.g., readmission outcome) generating summary statistics and basic data visualizations. Next, the development dataset, or raw data, is partitioned into 10 training and testing sets using stratified 10-fold cross-validation (CV). The remaining steps of Phases 1-3 are conducted separately within each of the 10 CV partitions. A standard scalar (from scikit-learn) is applied to transform features to remove the mean and scale to unit variance. This is important for certain ML algorithms to train optimally (i.e. logistic regression, K-nearest neighbors, and artificial neural networks).^9^ Next, any missing data values are imputed, since they are not allowed by most scikit-learn functions. Mode imputation is applied to categorical features, and subsequently an iterative imputer based on Multivariate imputation by chained equations (MICE)^10^ is applied to quantitative features. Notably, both the transformation and imputation are exclusively determined by the training data, and then similarly applied to each corresponding testing partition to avoid data leakage.

#### 3.3.2 Phase 2: Feature Importance and Feature Selection

Phase 2 evaluates feature importance within each CV partition using mutual information (MI) (proficient at detecting univariate associations)^11^, and MultiSURF (proficient at also detecting feature interactions). Features are selected using a simple collective feature selection approach, that only drops features identified as being uninformative(i.e. score *≤* 1) by both MI and MultiSURF prior to modeling.^12^ Because feature selection is conducted within each partition a different set of features may be passed to the next phase; however, this is also important for avoiding bias in the final testing evaluation.

#### 3.3.3 Phase 3: Machine Learning Modeling

Phase 3 conducts ML modeling applying 11 established classifiers each with their own strengths and weaknesses. Included are two simple classifiers; naive bayes^13^ and logistic regression^14^ that recognize simple univariate associations, but not epistatic interactions. Also included are five increasingly sophisticated tree-based classifiers including decision tree^15^, random forest^16^, gradient boosting^17^, extreme gradient boosting (XGB)^18^, and light gradient boosting (LGB).^19^ From this set of classifiers, decision trees are regarded as yielding interpretable models while the others are not. Additionally, this pipeline includes support vector machine (SVM), (testing linear, polynomial, and radial basis function kernels)^20^, artificial neural networks (ANN) (with a maximum of three hidden layers)^21^, and k-nearest neighbors classifier (k-Neighbors).^22^ These algorithms have been recognized to detect complex associations, but can be computationally expensive in large feature or instance spaces, and are often not regarded as yielding interpretable models. The last included algorithm is ExSTraCS 2.0^23^, an evolutionary rule-based ML classifier that has been demonstrated to be sensitive to both epistatic and heterogeneous associations with outcome. Rule-based ML methods like ExSTraCS are computationally expensive but are also regarded as being able to yield interpretable models unlike other advanced ML approaches that can yield high predictive performance but are considered black-box models.

Modeling in Phase 3 is conducted separately within each CV training partition. First, an automated hyperparameter optimization sweep is conducted using the Optuna package^24^ by further partitioning the training data into 3-fold training/testing sets. Throughout this study model optimization is based on balanced accuracy as the primary evaluation metric.^23^ A broad range of relevant hyperparameter options and ranges for each algorithm are hard-coded and documented within the aforementioned code on GitHub. Exceptions include naive Bayes, which does not have any hyperparameter options, and ExSTraCS, which is computationally expensive, but has fairly reliable default run parameters. With best hyperparameters selected, an optimized model is then trained from the entire training partition for each algorithm. Next, feature importance estimates are calculated for each model using permutation importance, which is the decrease in a model score when a single feature value is randomly shuffled.^25^ Then, all models are evaluated include all standard classification metrics, e.g. balanced accuracy, F1-score, area under the curve (AUC) of receiver operating characteristic (AUROC) and precision-recall (AUPRC) plots, and average precision statistic (APS) for AUPRC plots. In this study, particular emphasis was placed on the AUPRC as well as APS for final evaluation given the class imbalance of our target data.

#### 3.3.4 Phase 3: Post Analysis

Phase 4 post-analysis integrates and compares results over all ML algorithms and CV partitions to facilitate interpretation and downstream validation analyses. Average CV model performance metrics are calculated and non-parametric statistical comparisons (i.e. Kruskal-Wallis one-way AOV and Mann-Whitney U-tests) are performed to identify best performing algorithms, as well as demonstrate performance differences when the pipeline is applied to different datasets, e.g., all vs. 48 hour features, and development vs. validation. This pipeline also generates novel composite feature importance bar plots that summarize ML feature importance estimates across all ML algorithms. Prior to visualization, respective feature importance scores from each algorithm are normalized between 0-1 and weighted by the balanced accuracy of the given model, such that better performing algorithms contribute higher feature importance in these plots. In this study, the above pipeline was applied to train models that discern encounters that resulted in 30-day re-admissions from encounters that did not. All trained models were evaluated using a held-out validation dataset upon which our results are focused below. All model results are reported as the average measure across 10-fold cross validation.

### 3.4 Comparing Performance to EPIC Readmission Risk Model

To determine the potential impact of our classifiers to existing readmission models and policy interventions at the ABBCI, we compared the performance of the most predictive models for the validation set (full and 48 hour features) to the Epic Systems Corporation (Epic) Cognitive Computing Model (set at 24% threshold; full versus 48 hours) on the validation set using recall, precision, negative predictive value, specificity, and AUROC. For each model (full vs 48 hour features), we report the proportion of patients flagged by each individual model and characterize the patients based on age, sex, race, and socioeconomic status.

## 4 Results

Of the 2,460 patients studied in the development set, 10.6% (n=260 patients) had a 30-day readmission. When comparing the populations based on 30-day readmission status, we observed statistically significant differences based on age (70-79), sex, marital status (married and life partner), and cancer types (unspecified, blood, lung, male/female reproductive, breast, and lip). Additional characteristics between the readmission and non-readmission cohorts can be found in **Table 2**.

**Table 2:** Characteristics between cohort populations in development set

| Category | Subcategory | Readmission | Non-readmission | P-value |
| --- | --- | --- | --- | --- |
| Population |  | 260 (10.6%) | 2200 (89.4%) | <.00001 |
| Age | 18-29 | 5 (1.9%) | 55 (2.5%) | .5552 |
|  | 30-39 | 7 (2.7%) | 104 (4.7%) | .14156 |
|  | 40-49 | 20 (7.7%) | 222 (10.1%) | .2187 |
|  | 50-59 | 40 (15.3%) | 417 (18.9%) | .15854 |
|  | 60-69 | 68 (26.1%) | 585 (26.6%) | .86502 |
|  | 70-79 | 80 (30.8%) | 452 (20.6%) | **.00016** |
|  | 80-89 | 35 (13.5%) | 311 (14.1%) | .79486 |
|  | 90+ | 5 (1.9%) | 52 (2.4%) | .61708 |
| Sex | Female | 123 (47.3%) | 1159 (54.8%) | **.02202** |
|  | Male | 137 (52.7%) | 1041 (45.2%) | **.02202** |

Table 2 – continued from previous page
| Category | subcategory | Readmission | Non-readmission | P-value |
| --- | --- | --- | --- | --- |
| Race | White | 233 (89.6%) | 1979 (90.0%) | .84148 |
|  | Black | 16 (6.2%) | 113 (5.1%) | .45326 |
|  | Other | 8 (3.1%) | 74 (3.3%) | .86502 |
|  | Asian | 2 (0.8%) | 21 (1.0%) | .75656 |
|  | Unknown | 1 (0.4%) | 13 (0.6%) | .68916 |
| Marital Status | Married | 139 (53.5%) | 1351 (61.4%) | **.0139** |
|  | Single | 47 (18.1%) | 360 (16.3%) | .4593 |
|  | Widowed | 42 (16.1%) | 289 (13.1%) | .18024 |
|  | Divorced | 27 (10.3%) | 165 (7.5%) | .11184 |
|  | Legally Separated | 2 (0.8%) | 25 (1.1%) | .65994 |
|  | Life Partner | 1 (0.4%) | 1 (0.0%) | **.00298** |
|  | Unknown | 2 (0.8%) | 9 (0.4%) | .35758 |
| Social Determinants | Mean zip income | \$57,164.52 | \$57,668.41 | |
|  | Mean miles from hospital | 13.1 | 15 |  |
|  | Has financial concerns | 29 (11.2%) | 183 (8.3%) | .1141 |
| Cancer | Unspecified | 49 (24.5%) | 277 (19.2%) | **.04236** |
|  | Digestive | 57 (28.5%) | 338 (23.5%) | .07508 |
|  | Blood | 37 (18.5%) | 182 (12.6%) | **.00782** |
|  | Lung | 36 (18.0%) | 185 (12.8%) | **.0198** |
|  | Urinary | 19 (9.5%) | 126 (8.8%) | .70394 |
|  | Male reproductive | 17 (8.5%) | 236 (16.4%) | **.0009** |
|  | Breast | 15 (7.5%) | 238 (16.5%) | **.00016** |
|  | Female reproductive | 7 (3.5%) | 79 (5.5%) | **.17384** |
|  | Brain | 5 (2.5%) | 33 (2.3%) | .84148 |
|  | Endocrine | 3 (1.5%) | 20 (1.4%) | .89656 |
|  | Skin | 3 (1.5%) | 19 (1.3%) | .78716 |
|  | Soft Tissue | 3 (1.5%) | 18 (1.3%) | .78716 |
|  | Bone | 1 (0.5%) | 6 (0.4%) | .81034 |
|  | Non-malignant | 1 (0.5%) | 5 (0.3%) | .5892 |
|  | Lip | 0 (0.0%) | 22 (1.5%) | **.0466** |
| Cancer Counts | 1 | 151 (75.5%) | 1131 (78.5%) | .267 |
|  | 2 | 45 (22.5%) | 277 (19.2%) | .20408 |
|  | 3+ | 4 (2.0%) | 32 (2.3%) | .92034 |

### 4.1 Predicting 30-Day Readmissions

First, we assessed each 30-day readmission classifier’s performance in terms of area under the curve (AUROC) and precision/recall curve (AUPRC) leveraging all features versus features only available within the first 48 hours across development and validation sets below. Performance between the development and validation sets were comparable (**Table 5 in Supplement**). On the validation set, in **Figure 2**, among the classifiers trained using all features available, the highest AUROC was achieved by LGB (0.711) followed by Random Forest (0.707) and XGBoost (0.703). In terms of AUPRC, the highest APS is LGB (0.225), followed by XGBoost (0.212), and Random Forest (0.206), shown in **Figure 3**. Similarly, in **Figure 4**, among the classifiers trained using features available in the first 48 hours of admission, the highest AUROC was achieved by Random Forest (0.684) followed by XGBoost (0.681) and LGB (0.669). In terms of AUPRC, the highest APS was Random Forest (0.184) followed by XGBoost (0.182) and LGB (0.177) shown in **Figure 5**. According to the Mann Whitney test of significance, the Naive Bayes classifier was the only algorithm for which the performance between the model using all features compared to features available in the first 48 hours statistically was significant (AUROC p=0.032; APS p=0.0001).

**Figure 2:**
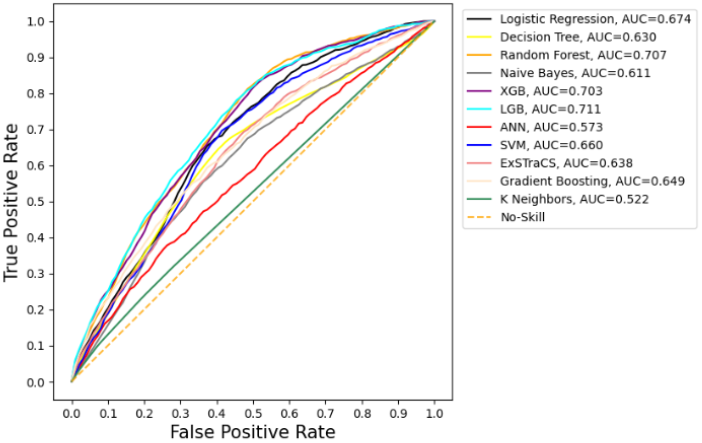
AUROC, all features, validation set

**Figure 3:**
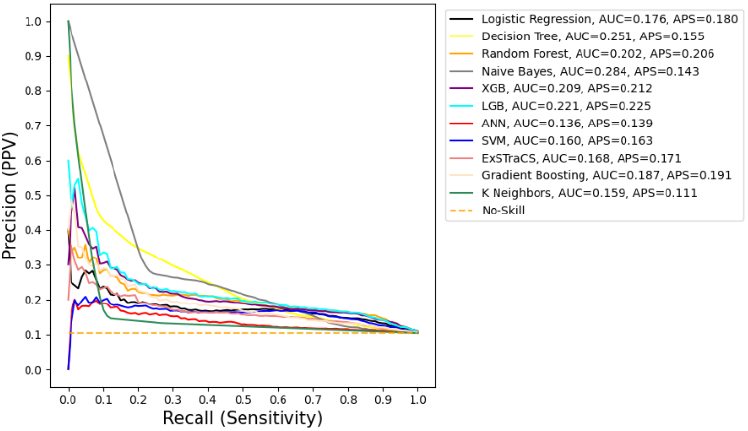
AUPRC, all features, validation set

**Figure 4:**
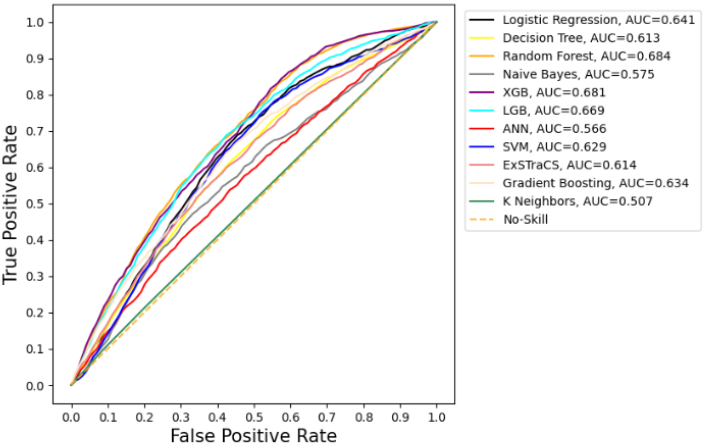
AUROC, 48-hr features, validation set

**Figure 5:**
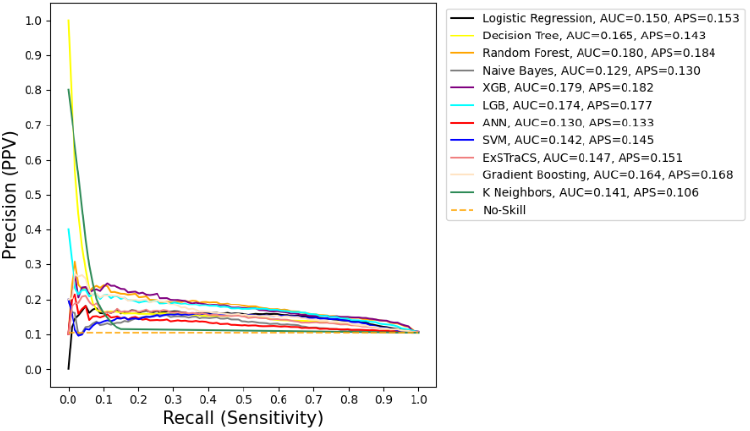
AUPRC, 48-hr features, validation set

### 4.2 Learning Feature Importance

We compared the most informative features for predicting 30-day readmission normalized and weighted across all algorithms using all features (**Figure 6**) compared to 48-hour features (**Figure 7**). Among all possible features, the most informative features include the LACE score and its individual components, laboratory values (sodium, albumin, hemoglobin), number of prior hospital admissions, time of discharge (epoch and winter), medical versus surgical population, admission type (emergency), Elixhauser score, length of stay, weight change over the past year, and cancer types of blood and digestive. The LACE score and last sodium and albumin values are among the most highly weighted.

**Figure 6:**
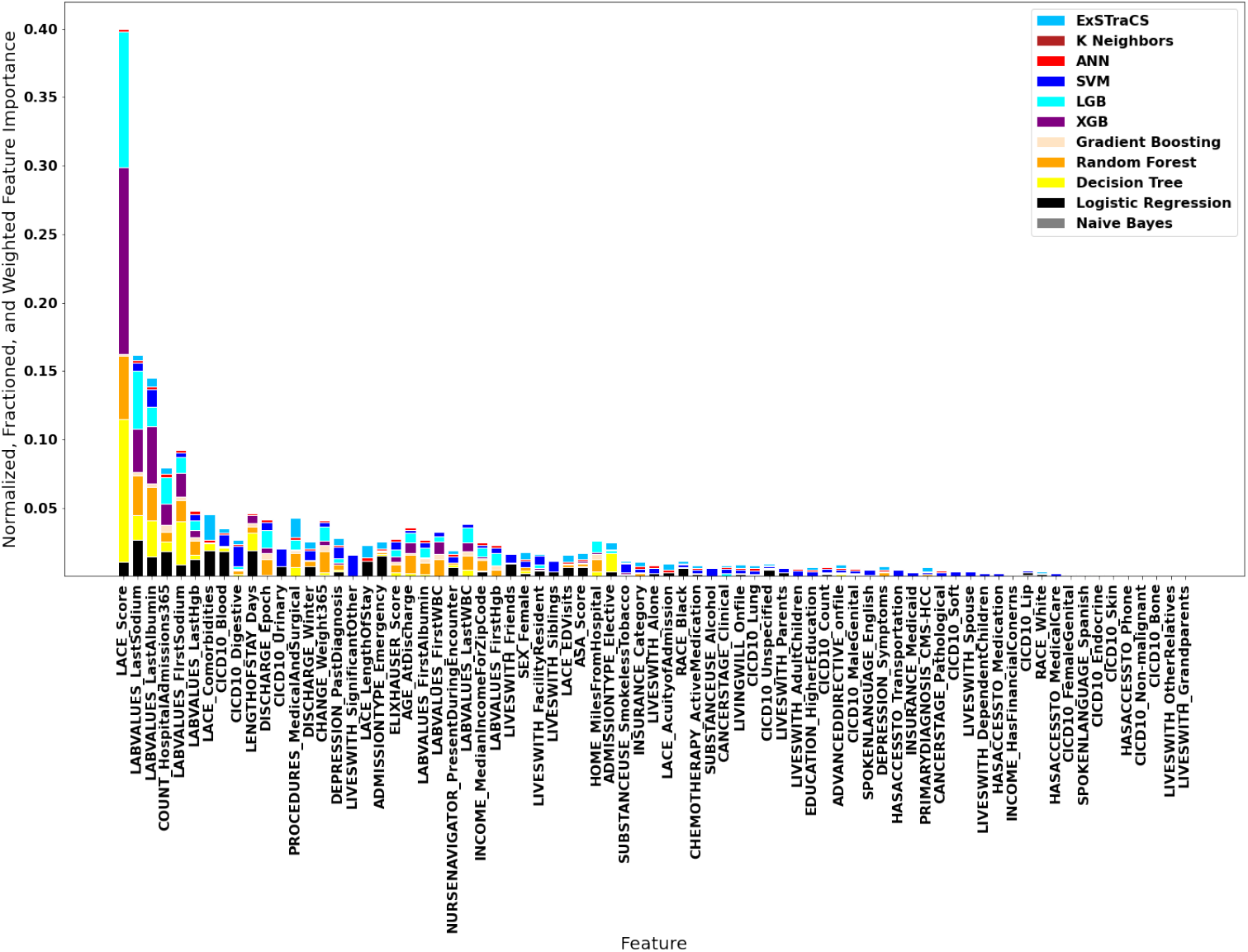
Plots of feature importance for all features

**Figure 7:**
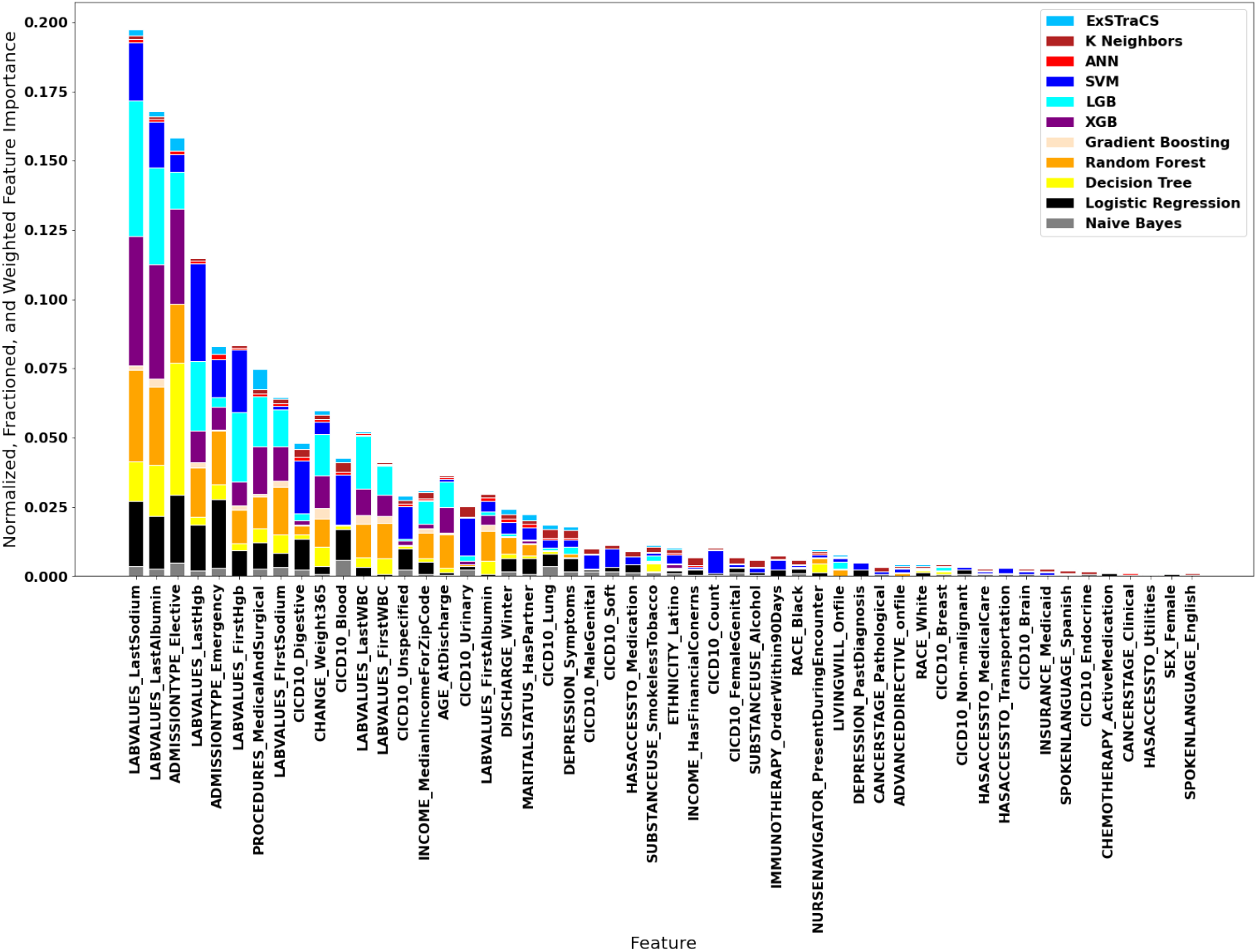
Plots of feature importance for 48-hr features

Similarities in the most important features of the 48-hour model included laboratory values (sodium, albumin, hemoglobin), time of discharge (winter), medical versus surgical population, admission type, weight change over the last year, and cancer types. However, the 48-hour model also contained novel community-level features including zip income and marital status, as well as additional cancer types (urinary, blood, unspecified), admission type (elective), and symptoms of depression.

We reviewed the most informative features for predicting 30-day readmission in the first 48 hours of admission among the most predictive models including random forest, LGB, and XGBoost. The most informative features across all models included laboratory test outcomes (sodium, albumin, hemoglobin, and white blood cells), hospital admission types (elective and emergency), change in weight over the past 365 days, medical surgical procedures, and age. Consistently least informative features include transportation services, utilities access, tobacco smoking status, and various types of cancer (skin, bone, lip, non-malignant, endocrine, and soft tissue cancers). Cancers with moderate informativeness include digestive, blood, and lung cancers.

### 4.3 Comparing Performance to Epic Readmission Risk Model

We compared our highest performing readmission risk model to the Epic model applied to all features and those only available within the first 48 hours of the encounter in **Table 3**. Leveraging all features, the LGB model produced higher, but comparable AUROC and APS compared to Epic. Given features in the first 48-hours, the random forest model produces higher AUROC, but lower AUPRC and APS than the Epic model.

**Table 3:** 30-day readmission classification on the validation set.

| Full dataset | AUROC | AUPRC | APS |
| --- | --- | --- | --- |
| LGB | 0.711 | 0.221 | 0.225 |
| Epic | 0.697 | 0.221 | 0.221 |
| 48-hr dataset | AUROC | AUPRC | APS |
| Random Forest | 0.684 | 0.180 | 0.184 |
| Epic | 0.676 | 0.197 | 0.198 |

**Table 4:** Characterization to assess diversity, equity, and inclusion among flagged patients by classifier for full validation set. RF=Random Forest

| Category | Subcategory | Full |  | 48-hour |  |
| --- | --- | --- | --- | --- | --- |
|  |  | LGB | EPIC | RF | EPIC |
| Age | 18-29 | 4 (1.1%) | 0 (0.0%) | 3 (0.6%) | 0 (0.0%) |
|  | 30-39 | 5 (1.3%) | 0 (0.0%) | 8 (1.8%) | 0 (0.0%) |
|  | 40-49 | 19 (5.0%) | 3 (3.6%) | 18 (4.1%) | 2 (3.6%) |
|  | 50-59 | 40 (10.6%) | 12 (14.5%) | 54 (12.3%) | 8 (14.3%) |
|  | 60-69 | 96 (25.3%) | 25 (30.1%) | 111 (25.2%) | 17 (30.3%) |
|  | 70-79 | 113 (29.8%) | 25 (30.1%) | 128 (29.1%) | 15 (26.8%) |
|  | 80-89 | 86 (22.7%) | 16 (19.3%) | 104 (23.6%) | 13 (23.2%) |
|  | 90+ | 16 (4.2%) | 2 (2.4%) | 14 (3.2%) | 1 (1.8%) |
| Sex | Female | 195 (51.3%) | 44 (53.0%) | 225 (51.1%) | 24 (42.9%) |
|  | Male | 185 (48.7%) | 39 (47.0%) | 215 (48.9%) | 32 (57.1%) |
| Race | White | 358 (88.1%) | 70 (84.3%) | 384 (87.3%) | 47 (83.9%) |
|  | Black | 22 (5.4%) | 6 (7.2%) | 27 (6.1%) | 3 (5.4%) |
|  | Other | 26 (6.5%) | 7 (8.4%) | 28 (6.6%) | 6 (10.7%) |
| Zip income | Average | \$58,321 | \$55,453 | \$58,787 | \$57,657 |
| Patients | Flagged as at risk | 380 (29.5%) | 83 (6.4%) | 440 (34.1%) | 56 (4.3%) |

**Table 5:** Characteristics between Cohort Populations

| Category | Subcategory | Development | Validation | P-value |
| --- | --- | --- | --- | --- |
| Population |  | 2460 (65.6%) | 1289 (34.4%) |  |
| Age | 18-29 | 60 (2.4%) | 31 (2.4%) | 1.00 |
|  | 30-39 | 111 (4.5%) | 57 (4.4%) | .9681 |
|  | 40-49 | 242 (9.8%) | 129 (10.0%) | .84148 |
|  | 50-59 | 457 (15.3%) | 211 (16.4%) | .37886 |
|  | 60-69 | 653 (26.5%) | 346 (26.8%) | .84148 |
|  | 70-79 | 532 (21.6%) | 293 (22.7%) | .4413 |
|  | 80-89 | 346 (14.1%) | 185 (14.4%) | .80258 |
|  | 90+ | 57 (2.3%) | 52 (2.8%) | .77182 |
| Sex | Female | 1282 (52.1%) | 706 (54.8%) | .11642 |
|  | Male | 1178 (47.9%) | 583 (45.2%) | .11642 |
| Race | White | 2212 (89.9%) | 1139 (88.4%) | .1556 |
|  | Black | 129 (5.2%) | 68 (5.3%) | .9681 |
|  | Other | 119 (4.8%) | 82 (6.4%) | .52218 |
| Marital Status | Married | 1490 (60.6%) | 772 (59.8%) | .63122 |
|  | Single | 407 (16.5%) | 217 (16.8%) | .8181 |
|  | Widowed | 331 (13.5%) | 172 (13.3%) | .86502 |
|  | Divorced | 192 (7.8%) | 101 (7.8%) | 1.0 |
|  | Legally Separated | 27 (1.1%) | 22 (1.7%) | .12356 |
|  | Unknown | 11 (0.4%) | 4 (0.3%) | .63122 |
|  | Life Partner | 2 (0.1%) | 1 (0.1%) | 1.0 |
| Social Determinants | Mean zip income | \$57,615.13 | \$57,544.38 | |
|  | Mean miles from hospital | 14.8 | 9.1 |  |
|  | Has financial concerns | 212 (8.6%) | 94 (7.3%) | .16758 |
| Cancer | Digestive | 395 (16.1%) | 156 (12.1%) | **.00104** |
|  | Mgen | 253 (10.3%) | 133 (10.3%) | 1.0 |
|  | Breast | 253 (10.3%) | 107 (8.3%) | **.04884** |
|  | Lung | 221 (8.9%) | 100 (7.7%) | .2113 |
|  | Blood | 219 (8.9%) | 80 (6.2%) | **.00374** |
|  | Other | 370 (15.0%) | 240 (18.6%) | **.00452** |
|  | Unspecified | 326 (13.3%) | 124 (9.6%) | **.00094** |
| Cancer Counts | 1 | 1282 (52.1%) | 617 (47.9%) | **.01468** |
|  | 2 | 322 (13.1%) | 139 (10.8%) | **.04136** |
|  | 3+ | 36 (1.4%) | 15 (1.2%) | .87288 |

**Table 6:** Features Used in the Prediction Model

| Variable Name | Variable Type |
| --- | --- |
| IMMUNOTHERAPY_OrderWithin90Days | binary |
| CHEMOTHERAPY_ActiveMedication | binary |
| DISCHARGE_Epoch | quantitative |
| DISCHARGE_Winter | binary |
| SEX_Female | binary |
| EDUCATION_HigherEducation | binary |
| SPOKENLANGUAGE_English | binary |
| SPOKENLANGUAGE_Spanish | binary |
| RACE_White | binary |
| RACE_Black | binary |
| ETHNICITY_Latino | binary |
| INSURANCE_Medicaid | binary |
| MARITALSTATUS_HasPartner | binary |
| ADMISSIONTYPE_Elective | binary |
| ADMISSIONTYPE_Emergency | binary |
| INCOME_MedianIncomeForZipCode | quantitative_continuous |
| LABVALUES_FirstAlbumin | quantitative_continuous |
| LABVALUES_LastAlbumin | quantitative_continuous |
| LABVALUES_FirstSodium | quantitative_continuous |
| LABVALUES_LastSodium | quantitative_continuous |
| LABVALUES_FirstWBC | quantitative_continuous |
| LABVALUES_LastWBC | quantitative_continuous |
| LACE_LengthOfStay | quantitative_discrete |
| LACE_AcuityofAdmission | quantitative_discrete |
| LACE_Comorbidities | quantitative_discrete |
| LACE_EDVisits | quantitative_discrete |
| LACE_Score | quantitative_discrete |
| INCOME_HasFinancialConcerns | binary |
| LENGTHOFSTAY_Days | binary |
| DEPRESSION_PastDiagnosis | binary |
| NURSENAVIGATOR_PresentDuringEncounter | binary |
| HASACCESSTO_Housing | binary |
| HASACCESSTO_Utilityies | binary |
| HASACCESSTO_Medication | binary |
| HASACCESSTO_Foods | binary |
| HASACCESSTO_MedicalCare | binary |
| HASACCESSTO_Transportation | binary |
| HASACCESSTO_Phone | binary |
| AGE_AtDischarge | quantitative_discrete |
| COUNT_HospitalAdmissions365 | quantitative_discrete |
| CHANGE_Weight365 | quantitative_continuous |
| ADVANCEDDIRECTIVE_onfile | binary |
| HOME_MilesFromHospital | quantitative_continuous |
| INSURANCE_Category | categorical_nominal |
| CANCERSTAGE_Clinical | categorical_ordinal |

**Table 6 – continued from previous page**
| **Variable Name** | **Variable Type** |
| --- | --- |
| CANCERSTAGE_Pathological | categorical_ordinal |
| PRIMARYDIAGNOSIS_CMS-HCC | binary |
| LIVESWITH_AdultChildren | binary |
| LIVESWITH_Alone | binary |
| LIVESWITH_DependentChildren | binary |
| LIVESWITH_FacilityResident | binary |
| LIVESWITH_Friends | binary |
| LIVESWITH_Grandparents | binary |
| LIVESWITH_OtherRelatives | binary |
| LIVESWITH_Parents | binary |
| LIVESWITH_Siblings | binary |
| LIVESWITH_SignificantOther | binary |
| LIVESWITH_Spouse | binary |
| ASA_Score | categorical_ordinal |
| DEPRESSION_Symptoms | binary |
| LIVINGWILL_Onfile | binary |
| LABVALUES_FirstHgb | quantitative_continuous |
| LABVALUES_LastHgb | quantitative_continuous |
| SUBSTANCEUSE_Alcohol | binary |
| SUBSTANCEUSE_SmokelessTobacco | binary |
| SUBSTANCEUSE_Tobacco | binary |
| ELIXHAUSER_Score | quantitative_discrete |
| PROCEDURES_MedicalAndSurgical | categorical_ordinal |
| CICD10_Lip | binary |
| CICD10_Digestive | binary |
| CICD10_Lung | binary |
| CICD10_Bone | binary |
| CICD10_Skin | binary |
| CICD10_Soft | binary |
| CICD10_Breast | binary |
| CICD10_FemaleGenital | binary |
| CICD10_MaleGenital | binary |
| CICD10_Urinary | binary |
| CICD10_Brain | binary |
| CICD10_Endocrine | binary |
| CICD10_Unspecified | binary |
| CICD10_Blood | binary |
| CICD10_Non-malignant | binary |
| CICD10_Count | quantitative_discrete |

In terms of the characteristics of patients flagged by these models, both the full (LGB) and 48-hour (random forest) feature models were highly sensitive in flagging more patients than the Epic models. Both models flagged patients with a similar distribution of race and sex; however, the LGB and random forest models flagged more patients among younger age groups. The Epic models were more sensitive to identifying patients with an average lower zip income.

## 5 Discussion

The goals of our study were to 1) compare prediction models using several ML algorithms to predict 30-day readmission among oncology patients using patient, provider, and community-level information available from the full visit versus first 48-hours, 2) learn the relative importance of risk factors across classifiers, and 3) determine the implications of our best model compared to existing models in the Epic EHR.

### 5.1 Predicting 30-Day Readmissions

The most predictive classifier, LGB, on the held-out validation set achieved an AUROC of 0.711 and APS of 0.225. Restricting our model to only features available within the first 48 hours, the best predictive classifier, a random forest, achieved an AUROC of 0.684 and APS of 0.184. Wong et al. classifiers (also gradient-boosted tree models with tree-explainers) performed with notably higher AUROC overall (0.78) and with features within first 24 hrs (0.74).^5^ We hypothesize that our gradient boosted tree models achieved lower predictive performance due to the simplicity of the features leveraged, e.g., omitting ICD-9 and LOINC-based embedding features capturing critical medical history. However, we determined that the performance of our 48-hour classifier (0.685) provides new insights into factors predictive of 30-day readmission.

### 5.2 Comparing Features for 30-day Readmission Risk Model

We observed several informative features predictive of readmission consistent in the literature including laboratory test outcomes (sodium, albumin, hemoglobin),^4^ hospital admission types (elective and emergency), discharge parameters,^4^ and age.^3^ Cancers with higher feature importance over other cancers include digestive, blood, and lung cancers.^3,4^ In contrast, many of predictive features in the literature were not as informative to our prediction models including race, having an advanced directive on file, insurance type, as well as demographics of race and sex. When comparing the features predictive of readmission within the first 24 hours described by Wong et al.^5^ and 48 hour models, we observed that blood cancer/hematology, age, and medical/surgical interventions received were common features.

### 5.3 Comparing Performance to Epic Readmission Risk Model

Finally, we compared our prediction models (full vs 48-hour) to the prediction model currently used in the LGH Epic EHR. In terms of full features, our LGB model performed with +1.4 points higher AUROC than the Epic model, but produced comparable AUPRC and APS results. In terms of the 48-hour model, our random forest model performed with +0.8 points higher AUROC than the Epic model, but produced slightly lower AUPRC (−1.7 points) and APS (−1.4 points) results. Although our results are comparable to the Epic model, our models diverged in important ways. Our 48-hour models were powered by novel features at various levels: patient (weight change over 365 days, depression symptoms, laboratory values, cancer type), provider (winter discharge, hospital admission type), community (zip income, marital status of partner). Our models provide several actionable insights to create service interventions deployed by the case management or discharge planning teams that may decrease readmission rates over time. For example, patients with large weight changes over the year or low albumin levels, could indicate issues with food scarcity, nutrition, and inability to ingest or keep down food. Providing patients with access to food programs/social workers, a nutritionist, or medications to control nausea and vomiting could mitigate 30-day readmissions with associated symptoms, respectively. Furthermore, patients experiencing depressive symptoms could be provided access to psychologists or a social support plan could be put into place. Patients with high white blood cell counts could have further evaluation to identify and mitigate cause as possible, including additional instruction for wound care and preventing infections. Patients flagged with risk of readmission may benefit from additional pre-discharge and ongoing outpatient education or be offered immediate access to tailored post-acute home care services. For patients who do not wish to receive post-acute care services in their homes, they could opt for frequent follow-up through phone calls or chatbots, e.g., send daily reminders for cleaning wounds, staying hydrated and well-fed, monitoring vital signs, and taking medications, etc.^26,27^ Combinations of clinical decision support and emerging digital health technologies may support transitions and provide the continuous care oncology patients need to reduce their readmission risk in the short-run. In **Figure 8**, we provide a prototypical representation of readmission risk model applied to a patient encounter with potential recommended action in Epic - figure adapted from Gallagher et al. 2020^28^ with additional approval from the Epic Corporation. We believe that leveraging Shapley values in clinical decision support systems can play a critical role in supporting explainable AI that can be readily interpreted and trusted to make informed clinical care decisions personalized to the patient’s circumstances and clinical status.

**Figure 8:**
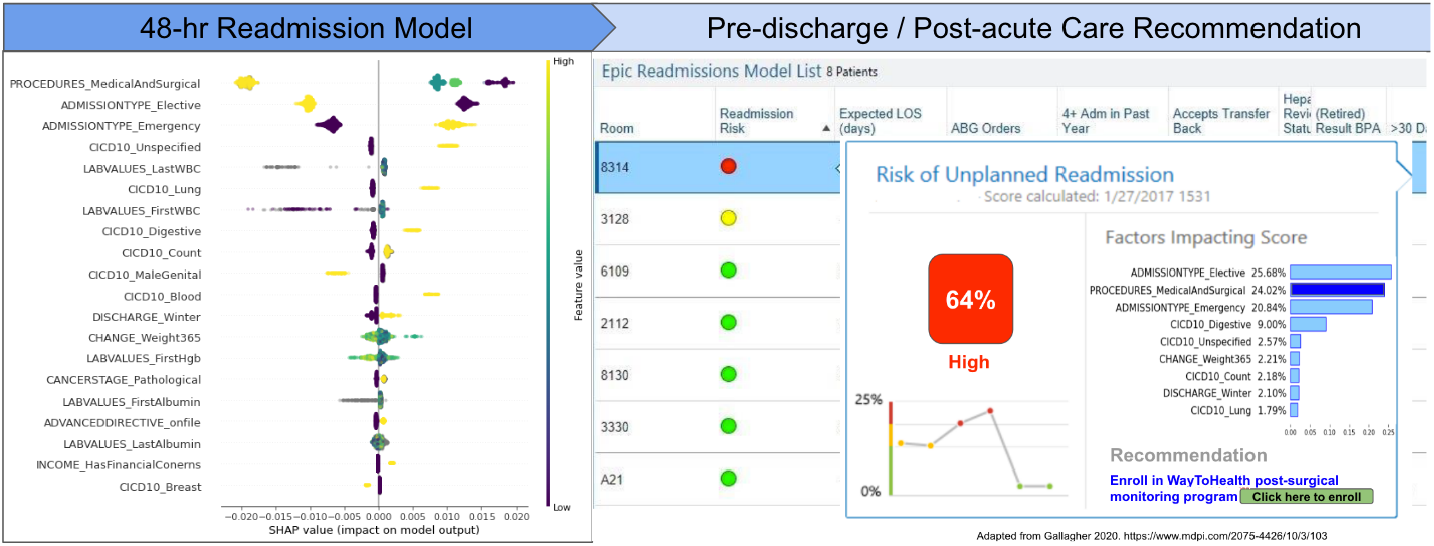
Translation representation of oncology prediction model (left) applied to current patient with potential recommended action (right).

In terms, of patients flagged by our model vs. the Epic model, both our full and 48-hour feature models were highly sensitive in flagging more patients than the Epic models. Given the large number of patients flagged, a prioritization scheme would be necessary to rank and identify those patients with the highest risk so as not to hinder clinical workflows. Both models flagged patients with a similar distribution of race and sex; however, our models flagged patients among younger age groups.

## 6 Limitations

Our project has several notable limitations. First, our prediction models were not evaluated at another hospital. We plan to test the portability of our models to our other Penn Medicine hospitals which utilize another implementation of the Epic EHR system. We also did not optimize the decision boundary for flagging a patient at risk for readmission instead opting for a default 0.50. Assessing the impact of this decision boundary could provide new opportunities to optimize the prediction model and potentially improve upon performance in future work.

## 7 Conclusions

In conclusion, we developed and validated robust 30-day readmission models for an oncology population. Our clinical findings have the potential for identifying oncology patients at risk for readmissions and the potential for implementing interventions that may impact patient care.

## Data Availability

The dataset is not available.

## 8 Acknowledgements

We thank Epic Corporation for their permission to include this prototypical representation of our oncology readmission risk model for their support.

## 9 Supplement

## 10 Keywords

ABBCI: Ann B. Barshinger Cancer Institute
ACP: Advanced Care Planning
ADL: Activities of Daily Living
ANN: Artificial Neural Network
APS: Average Precision Score
AUC: Area Under the Curve
AUPRC: Area Under the Precision Recall Curve
AUROC: Area Under the Receiver Operating Characteristic Curve
BMI: Body Mass Index
CV: Cross Validation
C4QI: Comprehensive Cancer Center Consoritums for Quality Improvement
ED: Emergency Department
EHR: Electronic Health Record
ExSTraCS: Extended Supervised Tracking and Classifying System
GI: Gastrointestinal
ICD-9 & 10: International Classification of Diseases, 9th and 10th revision
LGB: Light Gradient Boosting Machine
LGH: Lancaster General Hospital
LOINC: Logical Observation Identifiers Names and Codes
LOS: Length of Stay
ML: Machine Learning
MICE: Multivariate Imputation by Chained Equations
RF: Random Forest
SVM: Support Vector Machine
XGB: XGBoost
WBC: White Blood Cell Count

